# Mapping Structural Disconnection and Morphometric Similarity Alterations in Multiple Sclerosis

**DOI:** 10.1101/2024.06.19.24309154

**Authors:** Mario Tranfa, Maria Petracca, Marcello Moccia, Alessandra Scaravilli, Frederik Barkhof, Vincenzo Brescia Morra, Antonio Carotenuto, Sara Collorone, Andrea Elefante, Fabrizia Falco, Roberta Lanzillo, Luigi Lorenzini, Menno Schoonheim, Ahmed Toosy, Arturo Brunetti, Sirio Cocozza, Mario Quarantelli, Giuseppe Pontillo

**Affiliations:** Department of Advanced Biomedical Sciences, University “Federico II”, Naples, Italy; Department of Human Neurosciences, Sapienza University of Rome, Rome, Italy; Department of Molecular Medicine and Medical Biotechnology, Federico II University of Naples, Naples, Italy; Multiple Sclerosis Unit, Policlinico Federico II University Hospital, Via Sergio Pansini 5, 80131, Naples, Italy; Department of Neurosciences and Reproductive and Odontostomatological Sciences, University of Naples “Federico II”, Naples, Italy; Centre for Medical Image Computing, University College London, London, United Kingdom; Dementia Research Centre, UCL Queen Square Institute of Neurology, University College London, London, United Kingdom; Queen Square Multiple Sclerosis Centre, Department of Neuroinflammation, UCL Queen Square Institute of Neurology, University College London, London, United Kingdom; Department of Radiology and Nuclear Medicine, MS Center Amsterdam, Amsterdam Neuroscience, Amsterdam UMC, Vrije Universiteit Amsterdam, Amsterdam, The Netherlands; Department of Anatomy and Neurosciences, MS Center Amsterdam, Amsterdam Neuroscience, Amsterdam UMC, Vrije Universiteit Amsterdam, Amsterdam, The Netherlands; Institute of Biostructure and Bioimaging, National Research Council, Naples, Italy

**Keywords:** Multiple Sclerosis, structural disconnection, morphometric similarity, clinical disability, disability progression prediction, network-based statistics

## Abstract

Whilst multiple sclerosis (MS) can be conceptualized as a network disorder, brain network analyses are typically dependent on advanced MRI sequences not commonly acquired in clinical practice. Here, we used conventional MRI to assess cross-sectional and longitudinal modifications of structural disconnection and morphometric similarity networks in people with MS (pwMS), along with their relationship with clinical disability.

In this longitudinal monocentric study, 3T structural MRI scans of pwMS and healthy controls (HC) were retrospectively analysed. Physical and cognitive disabilities were assessed with the expanded disability status scale (EDSS) and the symbol digit modalities test (SDMT), respectively. Demyelinating lesions were automatically segmented on 3D-T1w and FLAIR images and, based on normative tractography data, the corresponding masks were used to compute pairwise structural disconnection between atlas-defined brain regions (100 cortical and 14 subcortical). Using the Morphometric Inverse Divergence (MIND) method, we built matrices of morphometric similarity between cortical regions based on FreeSurfer surface reconstruction. Using network-based statistics (NBS) and its prediction-based extension NBS-predict, we tested whether subject-level connectomes were associated with disease status, progression, clinical disability, and long-term confirmed disability progression (CDP), independently from global lesion burden and atrophy. The coupling between structural disconnection and morphometric similarity was assessed at different scales.

We studied 461 pwMS (age=37.2±10.6 years, F/M=324/137), corresponding to 1235 visits (mean follow-up time=1.9±2.0 years, range=0.1-13.3 years), and 55 HC (age=42.4±15.7 years; F/M=25/30). Long-term clinical follow-up was available for 285 pwMS (mean follow-up time=12.4±2.8 years), 127 of whom (44.6%) exhibited CDP. At baseline, structural disconnection in pwMS was mostly centered around the thalami and cortical sensory and association hubs, while morphometric similarity was extensively disrupted (*p*_FWE_<0.01). EDSS was related to fronto-thalamic disconnection (*p*_FWE_<0.01) and disrupted morphometric similarity around the left perisylvian cortex (*p*_FWE_ = 0.02), whilst SDMT was associated with cortico-subcortical disconnection in the left hemisphere (*p*_FWE_<0.01). Longitudinally, both structural disconnection and morphometric similarity disruption significantly progressed (*p*_FWE_ = 0.04 and *p*_FWE_<0.01), correlating with EDSS increase (rho 0.07, p 0.02 and rho 0.11, *p*<0.001), whilst baseline disconnection predicted long-term CDP with nearly 60% accuracy (*p* = 0.03). On average, structural disconnection and morphometric similarity were positively associated at both the edge (rho 0.18, *p*<0.001) and node (rho 0.16, *p*<0.001) levels.

Structural disconnection and morphometric similarity networks, as assessed through conventional MRI, are sensitive to MS-related brain damage and its progression. They explain disease-related clinical disability and predict its long-term evolution independently from global lesion burden and atrophy, potentially adding to established MRI measures as network-based biomarkers of disease severity and progression.

## Introduction

Multiple sclerosis (MS) is a chronic neuroinflammatory and neurodegenerative disease of the central nervous system, commonly associated with physical disability and cognitive impairment, and carrying an important personal and socio-economic burden.^1^ Whilst the assessment of focal lesions and, to some extent, brain atrophy using conventional MRI have key roles in the clinical management of MS, they only partially explain the clinical heterogeneity observed in people with MS (pwMS).^2^

From the field of network neuroscience, conceptualizing the brain as a complex system of gray matter (GM) regions - nodes - linked by structural and functional connections - edges, MS can be modeled as a network disorder.^3,4^ Demyelinating lesions disrupt white matter (WM) pathways connecting distant brain regions,^5^ while the development of atrophy subverts the ordered patterns of morphometric similarity between GM areas.^6^ Throughout MS, the accumulation of structural damage impacts the functional organization of the brain, ultimately leading to physical disability and cognitive impairment.^4^ Shifting the emphasis from characterizing damage in specific regions to understanding alterations at network level has yielded unprecedented insights into the pathophysiological mechanisms that underlie MS-related brain damage and associated clinical manifestations.^3^ However, brain network analyses are typically dependent on advanced MRI sequences that are not routinely acquired, hampering their implementation in clinical settings. This has led to considerable efforts in developing network analyses using anatomical images, to enable the (re-)analysis of conventional MRI datasets that were previously thought to lack network-level information.^7^

Structural disconnection between brain regions can be estimated from subject-level lesion masks, easily derived from anatomical images,^8^ and population-averaged tractography atlases, without requiring individual diffusion imaging.^9^ Such atlas-based approaches have demonstrated substantial agreement with individual tractography-based disconnectomes,^10^ offering an alternative perspective for evaluating the impact of MS lesions. Disconnection metrics correlate with physical disability^11^ and systemic biomarkers of axonal damage in MS,^12^ and disruption of specific brain subnetworks are linked to MS symptoms such as reduced information processing speed,^13^ memory dysfunction,^14^ or depression.^15^

Similarly, single-subject GM networks can be built from anatomical MRI by estimating a set of morphological properties (e.g., volume, thickness, curvature) within each GM region and computing the similarities between regions.^16^ Different methods utilizing this framework have demonstrated a restructuring of morphological similarity networks towards more disorganized configurations in pwMS, starting early in the disease,^17^ correlating with physical disability and cognitive impairment,^18^ and predicting disability worsening.^19^ However, these approaches have limitations, including how regions of interest are defined, the reliance on single metrics, or the reduction of complex data to simplistic summary statistics for each feature per region, making the link between GM networks and their neurobiological substrate somehow obscure.^16^ Recently, the Morphometric INverse Divergence (MIND) method has been proposed that addresses these limitations by estimating within-subject similarity between cortical areas based on the divergence between their multivariate distributions of multiple MRI features, with the advantages of higher technical reliability and biological validity.^20^

Most studies have assessed structural disconnection and morphometric similarity in isolation, using small sample sizes or short follow-up periods.^11,12,17,18^ Consequently, the potential of these measures as biomarkers of MS severity and progression remains largely unexplored. Here, leveraging a large monocentric cohort of pwMS, we jointly mapped structural disconnection and morphometric similarity both cross-sectionally and longitudinally. We aimed to demonstrate whether the corresponding networks: (i) are sensitive to MS-related brain damage and its progression over time; (ii) can explain MS-related physical disability and cognitive dysfunction; (iii) can predict long-term clinical worsening.

## Materials and methods

### Participants

In this retrospective longitudinal study, we analysed structural brain MRI and clinico-demographic data of patients with a diagnosis of MS according to the 2010-McDonald criteria^21^ and healthy controls (HC) from the radiological and clinical databases of the MS Center of the University of Naples “Federico II”. Exclusion criteria were age < 18 or > 75 years, and the presence of other relevant neurological, psychiatric, or systemic conditions. The study was conducted in compliance with the Declaration of Helsinki and approved by the Ethics Committee “Carlo Romano” of the Host Institution. Written informed consent was obtained from all participants.

### MRI acquisition

All MRI scans were acquired on the same 3T scanner (Magnetom Trio, Siemens Healthineers), equipped with an 8-channel head coil, from October 2006 to October 2020. The acquisition protocol included a 3D T1-weighted magnetization prepared rapid acquisition gradient echo sequence (MPRAGE - TR = 1900ms; TE = 3.4ms; TI = 900ms; flip angle 9°; voxel size 1 × 1 × 1 mm^3^; 160 axial slices) for morphometric analyses and, for pwMS, a T2-weighted Fluid Attenuated Inversion Recovery sequence (FLAIR - 3D: TR = 6000ms; TE = 396ms; TI = 2200ms; Flip Angle = 120°; voxel size 1×1×1 mm^3^; 160 sagittal slices; or 2D: TR = 9620ms; TE = 138ms; TI = 2500ms; Flip Angle = 150°; voxel size 1×1×3 mm^3^; 48 axial slices) for the assessment of demyelinating lesions.

### Clinical evaluation

Physical disability and information processing speed were assessed within one week from the MRI using the Expanded Disability Status Scale (EDSS) and the Symbol Digit Modalities Test (SDMT), respectively. SDMT values were converted into age-, sex- and education-adjusted z-scores based on normative values in the healthy population.^22^ For consistency with interpreting EDSS associations, SDMT z-scores were inverted before entering statistical analyses such that more positive values reflected poorer cognitive performance. When long-term (> 5 years) clinical follow-up was available, the EDSS was recorded and confirmed disability progression (CDP) with reference to the baseline examination was defined as an EDSS increase of ≥ 1 (for baseline EDSS ≤ 5.5) or ≥ 0.5 (for baseline EDSS > 5.5).^23^

### Lesion segmentation and structural disconnection networks

For all pwMS, demyelinating lesions were automatically segmented on FLAIR-T2w and T1w scans using the cross-sectional SAMSEG method in FreeSurfer v7.3.2.^24^ The obtained lesion masks were then used to compute total lesion volume (TLV) and to fill lesions in T1w images for subsequent morphometric analyses via FSL’s lesion filling procedure.^25^ Also, individual lesion masks were registered to the MNI template brain coordinate space by applying the nonlinear transformation obtained by normalizing each subject T1w volume to the template using ANTs v2.4.3.^26^ Spatially normalized lesion masks were used to obtain structural disconnection matrices based on a regional GM parcellation including 100 cortical regions from the Schaefer atlas^27^ and 14 subcortical regions from the FreeSurfer aseg atlas.^28^ Briefly, using the Lesion Quantification Toolkit,^29^ which relies on the HCP-842 diffusion MRI tractography atlas,^30^ the pairwise disconnection between structurally connected GM regions was computed as the proportion of streamlines intersecting lesions and used to fill subject-level 114 x 114 structural disconnection matrices. To help visualize the spatial distributions of lesions and resulting disconnection, we generated group-level lesion and disconnection probability maps, expressing the probabilities of each voxel containing a lesion or at least one streamline intersecting a lesion, respectively.^29^ To ease the visualization and interpretation of structural disconnection matrices, nodes were assigned to seven canonical functional system labels including visual (VIS), somatomotor (SM), dorsal attention (DAN), ventral attention (VAN), limbic (L), control (CONT), and default mode (DMN) networks,^31^ plus a network of subcortical regions (SUBC). Also, edge-level links were aggregated into region-level features by computing the sum of all values attached to each of the nodes.

### Structural MRI processing and morphometric similarity networks

Lesion-filled T1w volumes were processed with FreeSurfer v6.0.1 using the recon-all pipeline with default settings.^32^ As for lesion segmentation, different MRI visits were considered as separate instances using a cross-sectional image processing pipeline. Brain parenchymal fraction (BPF), considered a measure of global brain atrophy, was computed from FreeSurfer output as the ratio of brain volume to intracranial volume and expressed as z-scores adjusted for the effects of age and sex in healthy population. Based on FreeSurfer cortical surface reconstruction, vertex-level morphometric features (i.e., cortical thickness, GM volume, surface area, mean curvature and sulcal depth) were extracted for 100 regions of interest defined by the Schaefer atlas^27^ and used to compute the pairwise morphometric similarity between cortical regions using the MIND approach.^20^ Briefly, MRI features were standardized across all vertices and aggregated to form regional multivariate distributions. The similarity between each pair of regional multivariate distributions was computed based on the symmetrized Kullback-Leibler divergence metric and bounded between 0 and 1, with higher values representing greater similarity. The obtained values were used to fill subject-level 100 x 100 cortical morphometric similarity matrices. As for structural disconnection matrices, aggregated network- and region-level representations were also generated.

### Statistical analysis

Unless otherwise specified, statistical analyses were carried out using R (version 4.1.2). The effect of group (pwMS vs HC, only for morphometric similarity networks), EDSS, and SDMT scores on baseline structural disconnection and morphometric similarity networks were tested with the network-based statistics (NBS) approach,^33^ as implemented in the NBR package.^34^ NBS is a nonparametric method for performing statistical analysis on networks, that adjusts for multiple comparisons by clustering within topological rather than physical space. Briefly: 1) the hypothesis of interest is tested edge-wise using the general linear model; 2) connections are filtered according to a test statistic threshold; 3) connected graph components are identified among supra-threshold connections; 4) a family-wise error (FWE)-corrected p-value is computed for each component based on the sum of test statistic values using permutation testing.^33^ Likewise, longitudinal changes of structural disconnection and morphometric similarity networks were assessed using the implementation of linear mixed-effects models for NBS provided by the NBR package, with time points nested within subjects and random intercept and slope of follow-up time per subject. Similar mixed-effect models were used to assess the longitudinal evolutions of EDSS, TLV (log(x+1)-transformed to account for the positively skewed distribution), and z-scored BPF. For all NBS analyses, baseline age, age^2^ (to account for the nonlinear effect of age), and sex were included in the model as nuisance variables, with a primary statistical threshold of *p* < 0.01, 5000 permutations, and a statistical significance level set at *p*_FWE_ < 0.05. As we were interested in subnetwork-specific effects rather than the influence of global lesion burden or atrophy, models assessing the correlations with clinical variables were additionally adjusted for log-transformed TLV (for structural disconnection matrices) and BPF z-scores (for morphometric similarity matrices). Also, when subnetworks exhibiting significant change over time emerged, these were summarized for further analyses by z-scoring each edge using the healthy population as a reference and calculating the average of their modules to obtain global synthetic measures of longitudinal structural disconnection and morphometric similarity change. Specifically, we utilized Spearman’s rank correlation to test the associations of structural disconnection and morphometric similarity disruption over time with annualized EDSS change. Also, the longitudinal association between longitudinal structural disconnection and morphometric similarity disruption and disability worsening were reassessed while accounting for changes in log-transformed TLV and BPF z-scores, respectively, using partial correlations.

To evaluate the prognostic value of structural disconnection and morphometric similarity, we tested whether baseline networks could predict long-term CDP using the NBS-Predict approach, a prediction-based extension of NBS combining machine learning models with connected components in a cross-validation (CV) structure, as implemented in the corresponding MATLAB (MathWorks, 2017) toolbox.^35^ We ran NBS-Predict with 5-fold nested CV (primary threshold *p* < 0.01) and hyperparameter optimization using Bayesian optimization with 100 iterations. The CV structure was repeated ten times to reduce the variation in the model performance estimation. We scaled data and regressed out baseline age, age^2^, sex, and log-transformed TLV (for structural disconnection matrices) and BPF z-scores (for morphometric similarity matrices), using a cross-validated deconfounding technique to prevent data leakage.^36^ Different machine learning algorithms (logistic regression, linear support vector classification, and linear discriminant analysis) were evaluated, with classification accuracy as the performance metric and 500 permutations to assess the significance of the models’ predictions.^35^

Finally, to investigate the relationship between structural disconnection and morphometric similarity in pwMS at baseline, we utilized Spearman’s rank correlation to test the associations between the two networks at different scales. At the global network level, we determined the group-level correlation between mean structural disconnection and mean morphometric similarity across subjects. At the edge level, we first averaged structural disconnection and morphometric similarity networks across subjects and then correlated the two vectorized matrices. We also repeated this analysis using individual (rather than average) matrices to obtain a distribution of coupling values across subjects.^37^ At the node level, regional connectivity profiles were extracted from each row of the structural disconnection and morphometric similarity matrices and correlated with each other to obtain coupling values for each of the 100 cortical parcels.^38^

## Results

### Participants

We analysed a total of 461 pwMS (F/M = 324/137, mean age = 37.2 ± 10.6 years) corresponding to 1235 visits (median number of visits per patient 4, range 1 - 8; mean follow-up time = 1.9 ± 2.0 years, range = 0.1 - 13.3 years), and 55 healthy controls (F/M = 25/30, mean age = 42.4 ± 15.7 years).

Baseline EDSS (median = 2.5, interquartile range = 2.0 - 4.0) and SDMT (mean z-score = −1.1 ± 1.1) were available for 459 and 247 pwMS, respectively. Long-term clinical follow-up was available for 285 pwMS (mean follow-up time = 12.4 ± 2.8 years), 127 of whom (44.6%) exhibited CDP. Demographic, clinical, and MRI characteristics of the studied population are reported in Table 1.

**Table 1.**
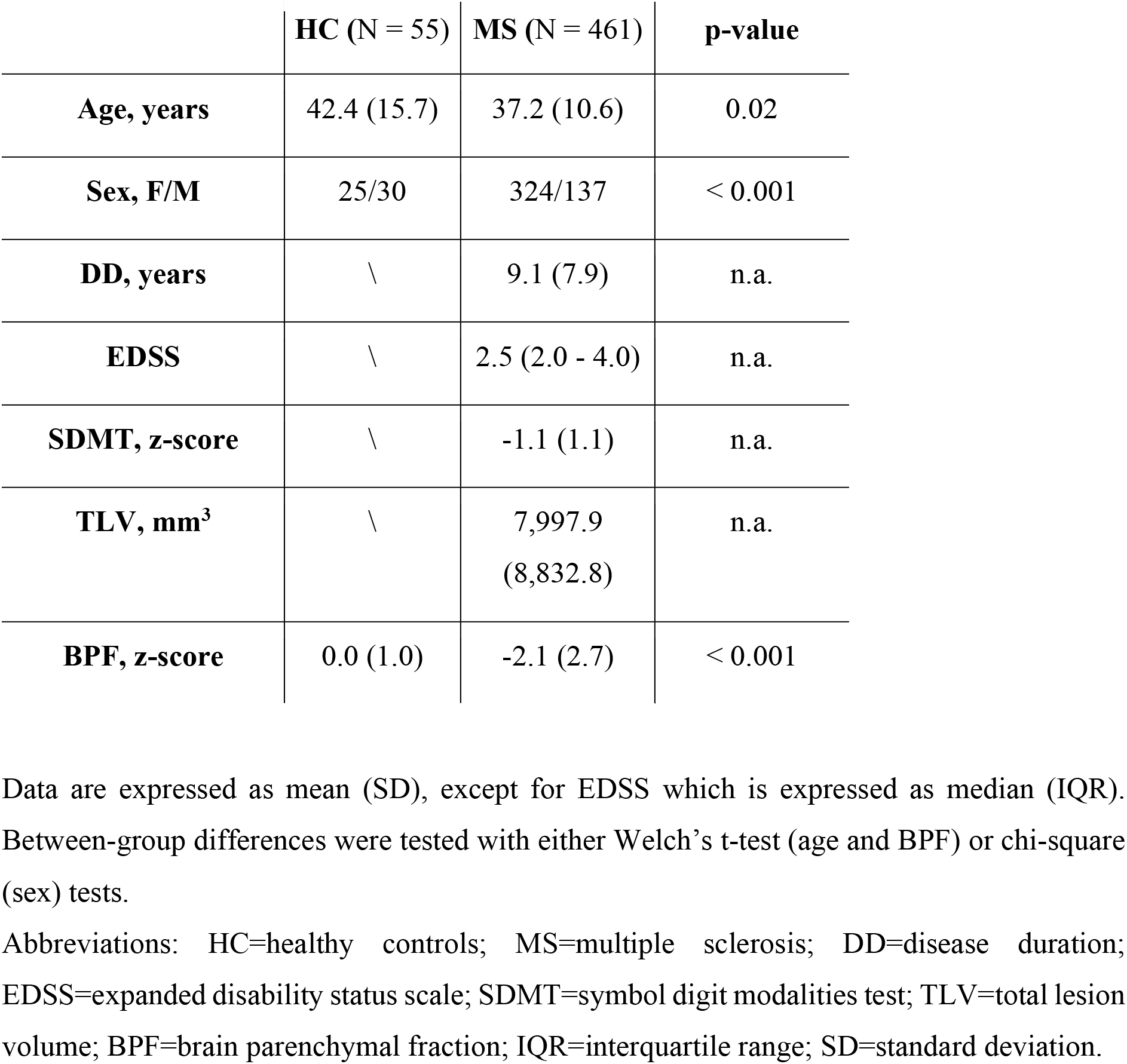
Demographical and clinical data of the study population.

|  | **HC (N = 55)** | **MS (N = 461)** | **p-value** |
| --- | --- | --- | --- |
| **Age, years** | 42.4 (15.7) | 37.2 (10.6) | 0.02 |
| **Sex, F/M** | 25/30 | 324/137 | < 0.001 |
| **DD, years** | \ | 9.1 (7.9) | n.a. |
| **EDSS** | \ | 2.5 (2.0 - 4.0) | n.a. |
| **SDMT, z-score** | \ | -1.1 (1.1) | n.a. |
| **TLV, mm<sup>3</sup>** | \ | 7,997.9<br>(8,832.8) | n.a. |
| **BPF, z-score** | 0.0 (1.0) | -2.1 (2.7) | < 0.001 |

Data are expressed as mean (SD), except for EDSS which is expressed as median (IQR). Between-group differences were tested with either Welch's t-test (age and BPF) or chi-square (sex) tests.
Abbreviations: HC=healthy controls; MS=multiple sclerosis; DD=disease duration; EDSS=expanded disability status scale; SDMT=symbol digit modalities test; TLV=total lesion volume; BPF=brain parenchymal fraction; IQR=interquartile range; SD=standard deviation.

### Structural disconnection and morphometric similarity disruption in MS

At baseline, pwMS displayed the highest lesion probability in the periventricular WM (Figure 1A), with the highest disconnection probability at the level of the occipital WM, splenial commissural fibers, and long-range frontal and temporal association tracts (Figure 1B). On average, structural disconnection was mainly observed between the VIS and the SM and non-sensorimotor networks, as well as within and between cortical association networks and around subcortical structures (Figure 1C). At the regional level, the most structurally disconnected nodes were the thalami and temporal and posterior cortical regions (Figure 1D).

**Figure 1.**
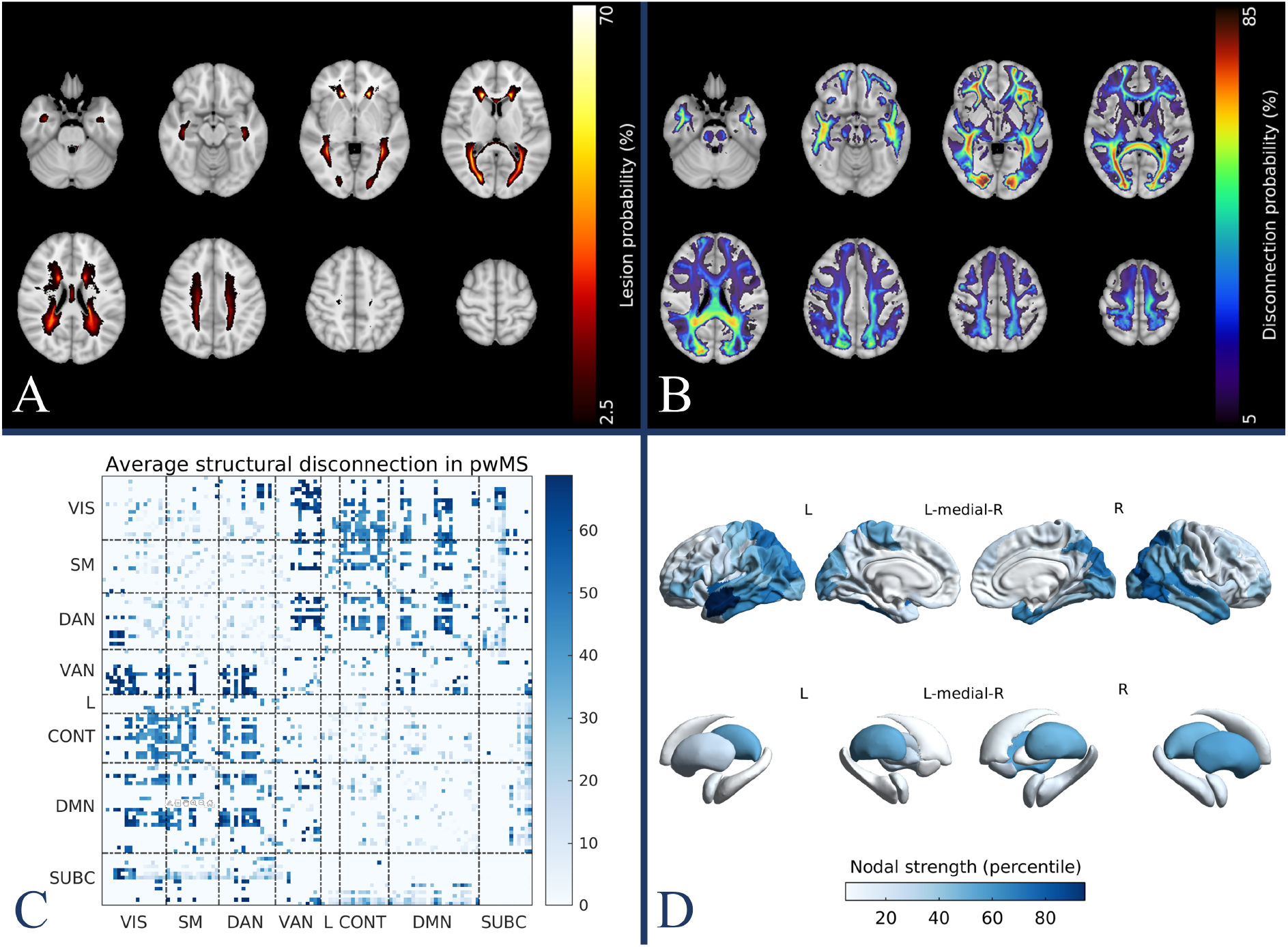
Structural disconnection in pwMS. Group-level lesion (A) and disconnection (B) probability maps in pwMS, expressing the probabilities of each voxel containing a lesion or at least one streamline intersecting a lesion, respectively. Network- (C) and region- (D) level representations of average structural disconnection in pwMS. *Abbreviations: pwMS patients with multiple sclerosis*.

Compared with HCs (Figure 2A), pwMS showed a distributed subnetwork of predominantly disrupted morphometric similarity (431 edges, *p*_FWE_ < 0.01, Figure 2B-C), with the prominent involvement of occipital, pericentral, perisylvian, and prefrontal cortices (Figure 2D). Global structural disconnection and morphometric similarity disruption significantly correlated with TLV (Spearman’s rho 0.94, *p* < 0.001, Supplementary Figure 1A) and BPF (Spearman’s rho −0.40, *p* < 0.001, Supplementary Figure 1B), respectively.

**Figure 2.**
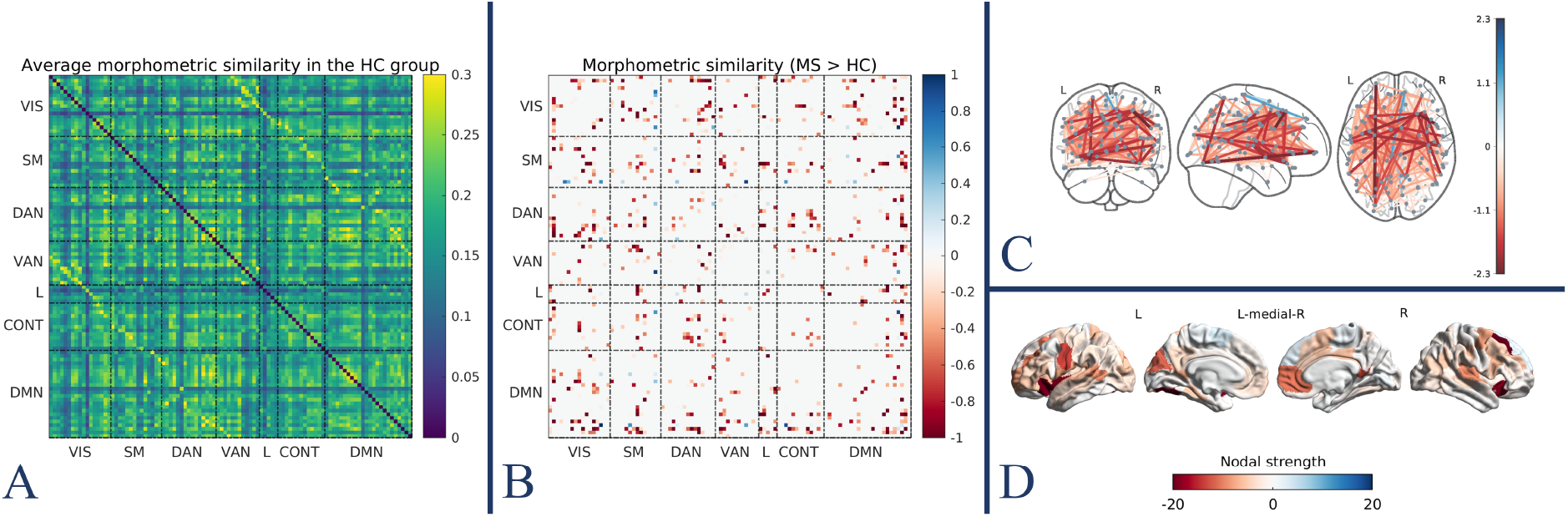
Morphometric similarity disruption in pwMS. Average morphometric similarity network in the HC group (A). Network- (B), edge- (C), and region- (D) level representations of the subnetwork of significant between-group differences in terms of morphometric similarity (pwMS > HC). *Abbreviations: pwMS=patients with multiple sclerosis; HC healthy controls*.

### Structural disconnection and morphometric similarity disruption explain physical and cognitive disability

At baseline, we found a subnetwork of significant association between EDSS and structural disconnection (225 edges, *p*_FWE_ < 0.01), mainly involving cortico-subcortical tracts, within-transmodal connections of the DMN, the DAN and the CONT, and links between these and sensorimotor networks. The regions participating the most in this subnetwork were the thalami, the amygdalae, and the prefrontal cortex (Figure 3A-C). EDSS was also significantly associated with a smaller subnetwork of predominantly disrupted morphometric similarity between the DAN and the other networks, with the prominent participation of the insula and the perisylvian cortex of the left hemisphere (86 edges, *p*_FWE_ = 0.02) (Figure 3D-F). Similarly, SDMT was associated with a relatively small subnetwork of predominantly cortico-subcortical structural disconnection mostly involving the left hemisphere (88 edges, *p*_FWE_ < 0.01), with the participation of the thalamus and prefrontal, temporal, and occipital cortical regions (Figure 4). No significant subnetworks emerged when assessing the relationship between morphometric similarity and SDMT.

**Figure 3.**
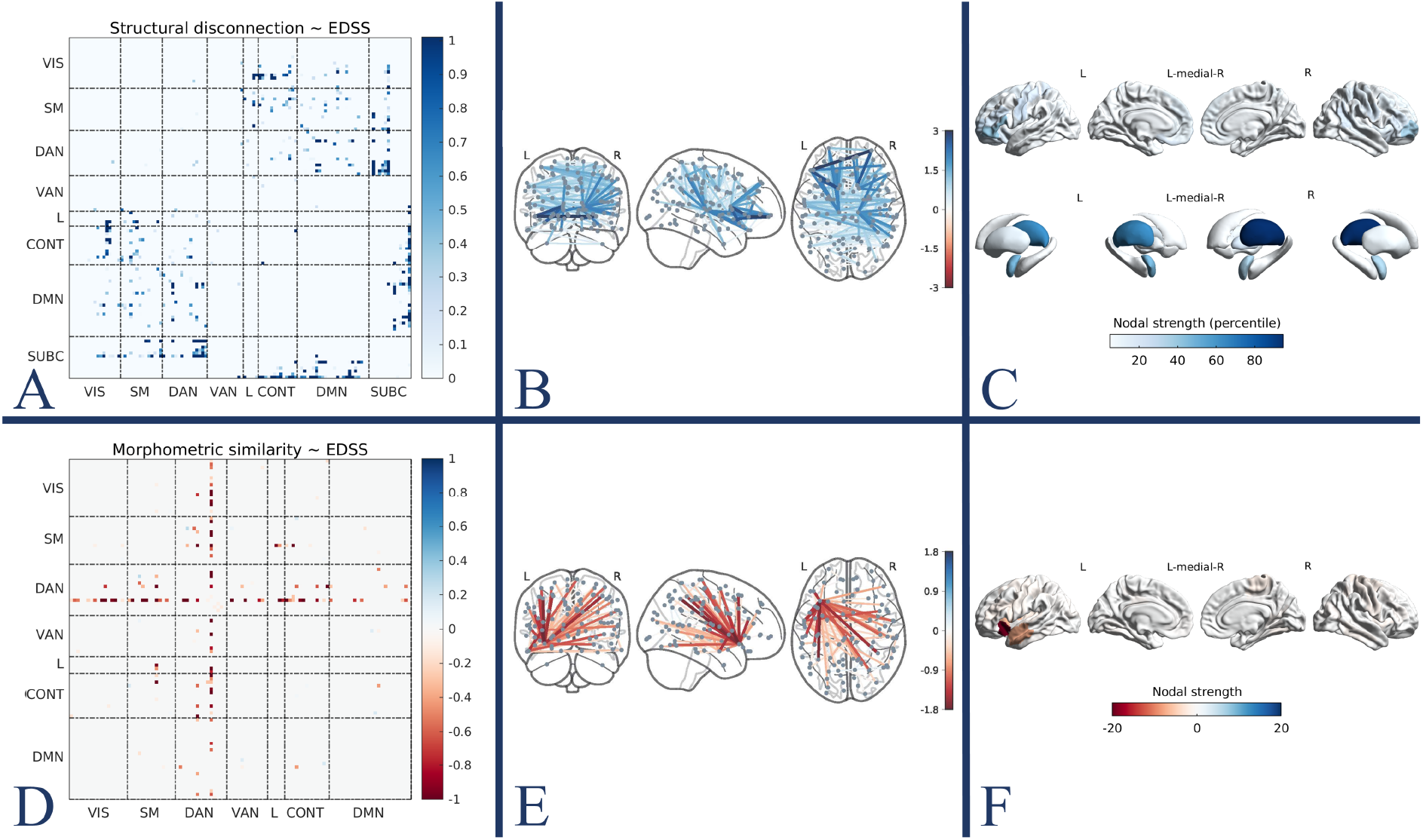
Structural disconnection and morphometric similarity disruption explain physical disability. Network- (A), edge- (B), and region- (C) level representations of the subnetwork of significant association between EDSS and structural disconnection. Network- (D), edge- (E), and region- (F) level representations of the subnetwork of significant association between EDSS and morphometric similarity. *Abbreviations: EDSS expanded disability status scale*.

**Figure 4.**
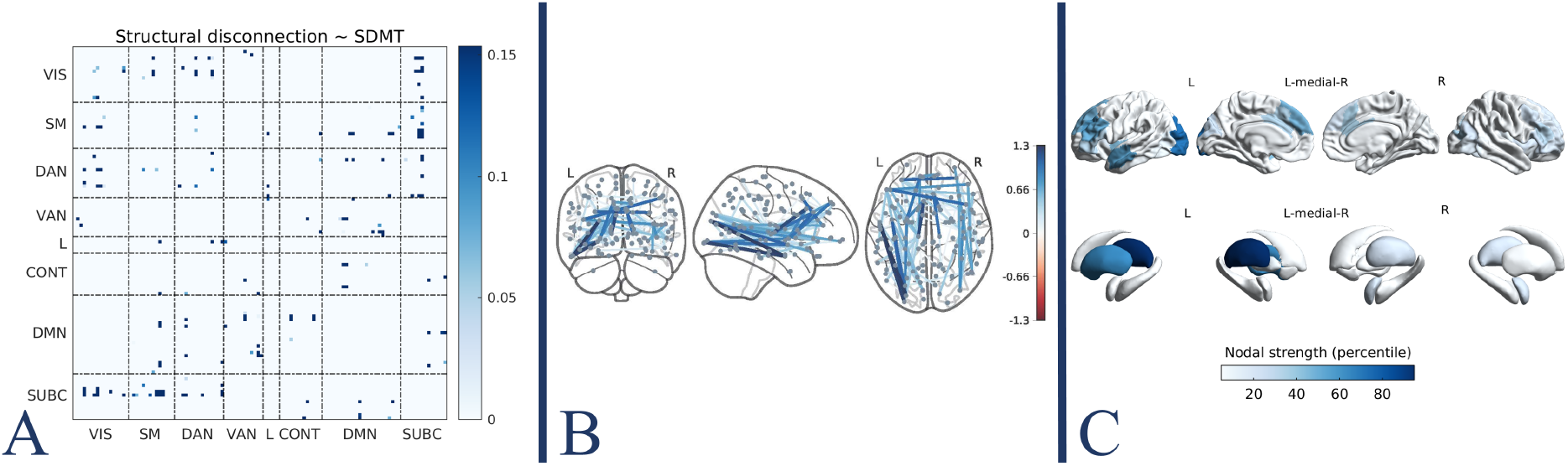
Structural disconnection explains cognitive performance. Network- (A), edge- (B), and region- (C) level representations of the subnetwork of significant association between SDMT and structural disconnection. *Abbreviations: SDMT symbol digit modalities test*.

### Structural disconnection and morphometric similarity disruption progress over time and explain disability worsening

Longitudinally, we found a smaller subnetwork of progressive structural disconnection (82 edges, *p*_FWE_ = 0.04) involving mainly fronto-thalamic tracts (Figure 5A-C). Moreover, we observed a larger and anatomically distributed subnetwork of progressive morphometric similarity alterations (509 edges, *p*_FWE_ < 0.01), comprising pairs of regions exhibiting both increased and decreased similarity over time (Figure 5D-F). Longitudinal models also showed significant EDSS worsening (*B* = 0.084, *SE B* = 0.015, *p* < 0.01), and whole-brain volume loss (BPF: *B* = −0.095, *SE B* = 0.022, *p* < 0.01) over time, while the increase in global lesion burden was not significant (logTLV: *B* = 0.003, *SE B* = 0.004, *p* = 0.50). Annualized structural disconnection significantly correlated with annualized changes in EDSS scores (Spearman’s rho 0.07, *p* = 0.02), with this correlation remaining significant after adjusting for longitudinal TLV change (Spearman’s rho 0.08, *p* = 0.004). Similarly, individualized morphometric similarity change per year correlated with annualized EDSS change (Spearman’s rho 0.11, *p* < 0.001), with this correlation remaining significant also after accounting for annualized BPF change (Spearman’s rho 0.09, *p* = 0.002).

**Figure 5.**
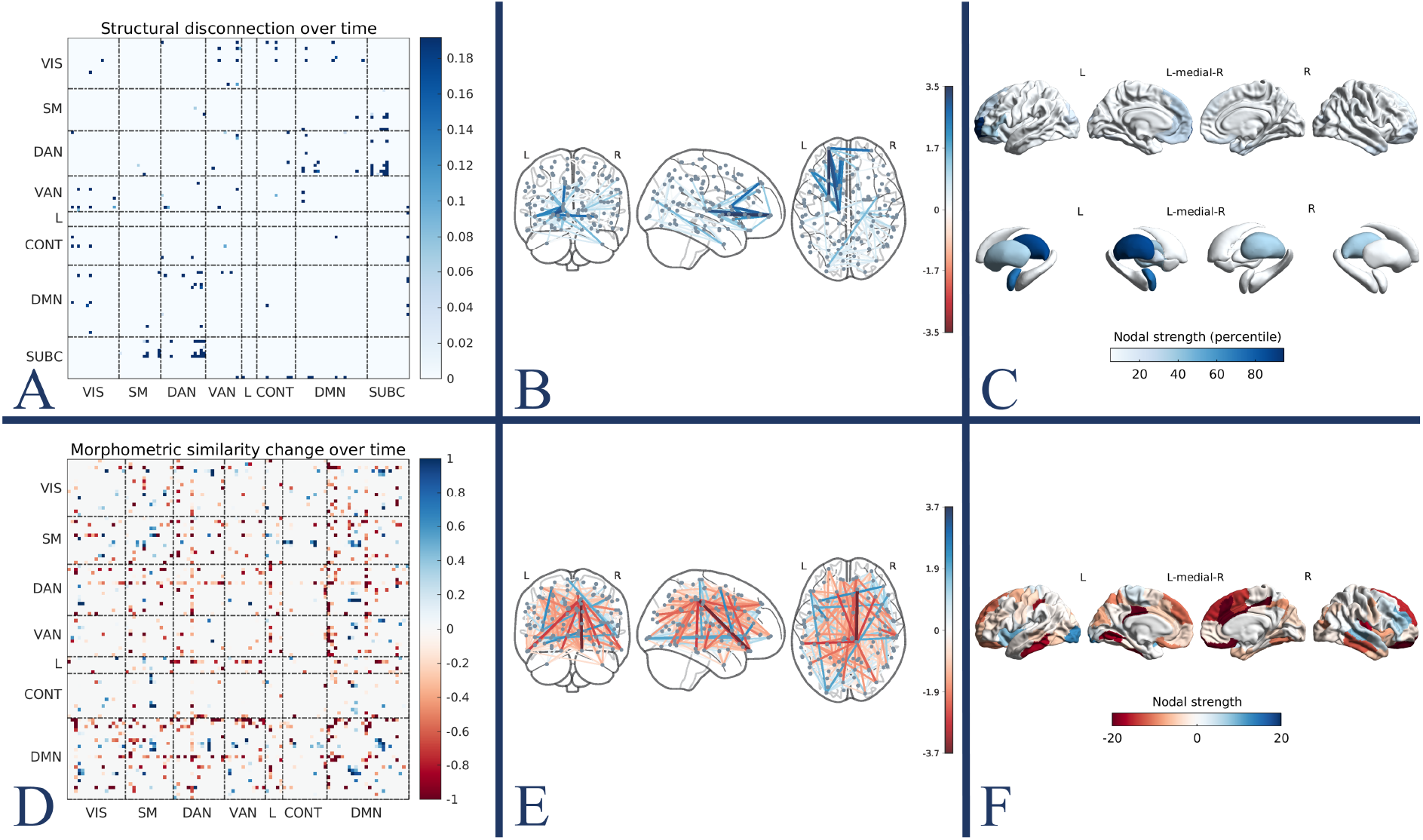
Structural disconnection and morphometric similarity changes over time. Network- (A), edge- (B), and region- (C) level representations of the subnetwork of significant structural disconnection over time in pwMS. Network- (D), edge- (E), and region- (F) level representations of the subnetwork of significant morphometric similarity change over time in pwMS. *Abbreviations: pwMS patients with multiple sclerosis*.

### Baseline structural disconnection predicts long-term disability progression

Using NBS-predict with the baseline structural disconnection matrices as input, a linear support vector machine classifier significantly predicted long-term CDP (710 edges, accuracy 0.59, _95%_CI 0.58 - 0.60, *p* = 0.03). The identified subnetwork mainly involved cortico-subcortical tracts, within-transmodal connections of the DMN, the DAN, the VAN, and the CONT, and links between these and sensorimotor networks. The regions participating the most in this subnetwork were preferentially located in the left hemisphere and included the thalamus and the parieto-occipital, pericentral, and prefrontal cortices (Figure 6). Models relying on baseline morphometric similarity matrices did not achieve above chance-level accuracy for the prediction of CDP.

**Figure 6.**
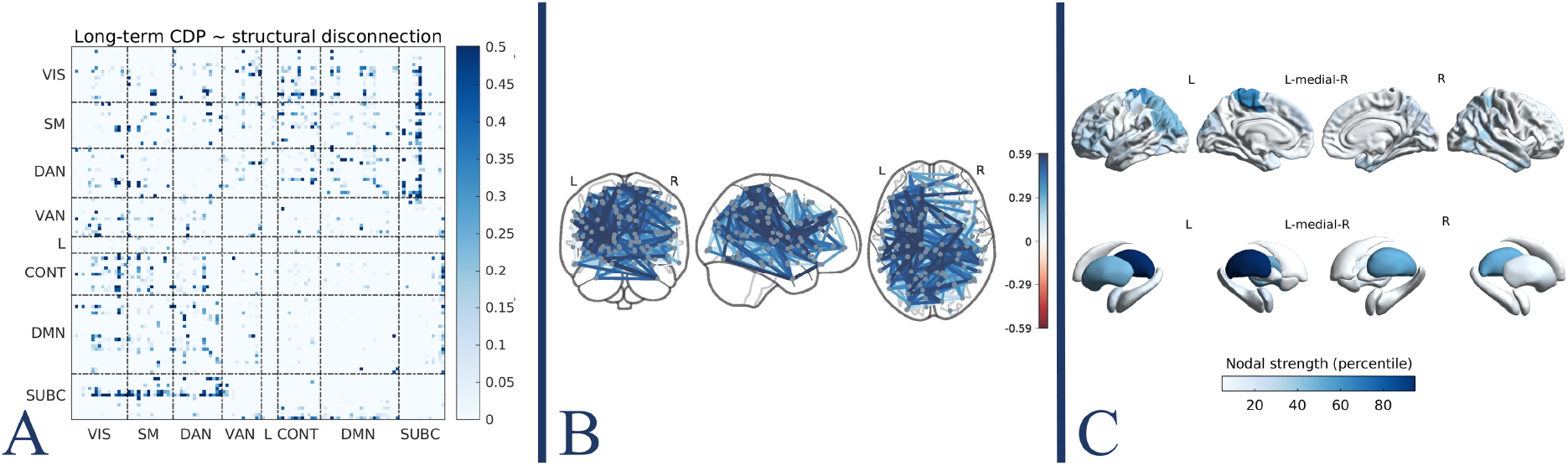
Baseline structural disconnection predicts long-term disability progression. Network- (A), edge- (B), and region- (C) level representations of the structural disconnection subnetwork predicting long-term CDP. *Abbreviations: CDP confirmed disability progression*.

### Coupling between structural disconnection and morphometric similarity

At the global network level, higher mean structural disconnection in pwMS was associated with lower mean morphometric similarity (Spearman’s rho −0.18, *p* < 0.001) (Figure 7A). Conversely, at a more granular level, edges with a greater probability of structural disconnection were generally associated with higher morphometric similarity (Spearman’s rho 0.18, *p* < 0.001), with the distribution of values suggesting a nonlinear, multiphasic, relationship between the two (Figure 7B and Supplementary Figure 2). Similarly, at the node level, there was a positive association between regional structural disconnection and morphometric similarity profiles (average Spearman’s rho 0.16, *p* < 0.001) with the strongest coupling observed at the level of sensorimotor areas and fronto-parietal association hubs (Figure 7C).

**Figure 7.**
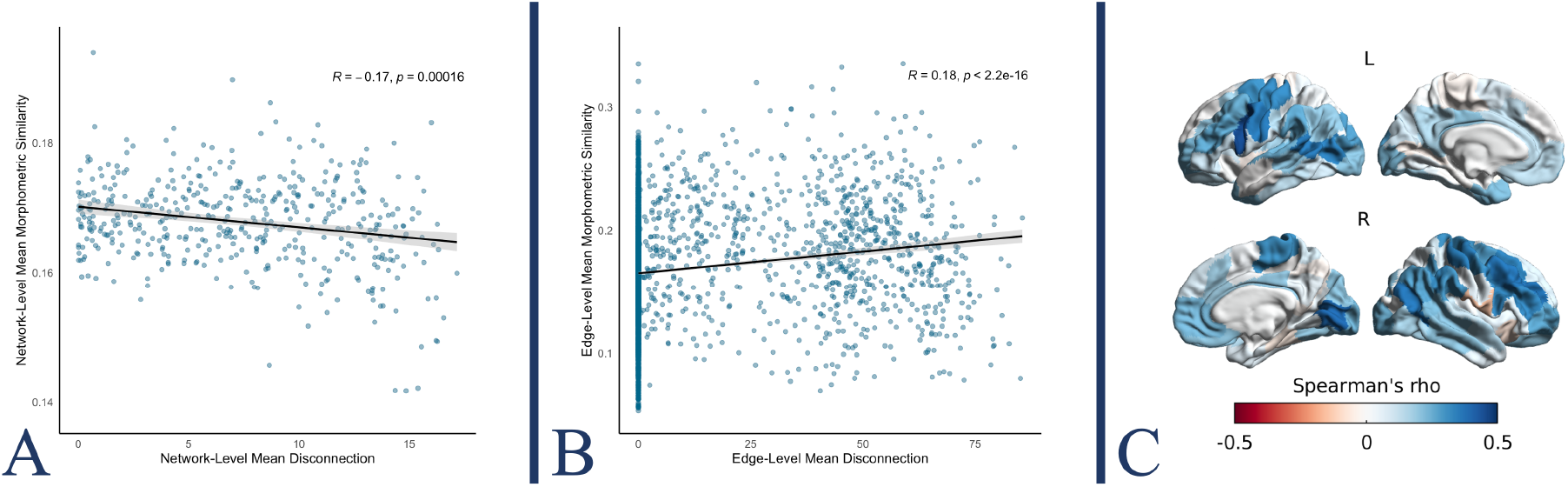
Coupling between structural disconnection and morphometric similarity. Network- (A), edge- (B), and region- (C) level associations between structural disconnection and morphometric similarity. (A) Group-level correlation between mean structural disconnection and mean morphometric similarity across subjects. (B) Edge-level correlation between average structural disconnection and morphometric similarity networks. (C) For each of the 100 cortical parcels, correlation between regional structural disconnection and morphometric similarity profiles.

## Discussion

Using network analysis methods and conventional MRI sequences, we identified patterns of structural disconnection and morphometric similarity disruption in the brains of pwMS. These measures proved to be sensitive to disease progression and were able to explain disease-related clinical disability and predict its long-term evolution, independently from global lesion burden and atrophy.

In line with well-established evidence,^39^ the highest T2 lesion occurrence in our cohort was observed in the bilateral periventricular WM, probably reflecting the preferential perivenular distribution of MS-related inflammatory demyelination.^40^ As the anatomical configurations of long-range and cortico-subcortical tracts make them more likely to traverse lesional areas, resulting in more severe damage, structural disconnection was mainly centered around the thalami and cortical sensory (occipital and pericentral cortices) and association (temporal cortex) hubs. The thalamus, in particular, with its multiple reciprocal connections, is sensitive to lesions occurring in many different regions, thus acting as a “barometer” for diffuse brain parenchymal damage.^41^ Likewise, the visual and sensorimotor cortices, as well as the temporal cortex, are served by long-range tracts sustaining their function as primary sensory areas and integration hubs, respectively, thus being particularly prone to structural disconnection.^42,43^ While TLV and average structural disconnection are linked by a logarithmic relationship that is enforced by the brain’s geometry, not all lesion locations bear equal clinical relevance, differentially affecting physical disability,^44^ cognition,^45^, and long-term clinical outcomes.^46^ The examination of topologically distributed effects, using structural disconnection networks, complements the impact of MS lesions, potentially resulting in more robust neurobiological and clinical associations compared with the assessment of isolated regions/connections.^47,48^ Indeed, we found that, independent of the global lesion burden, physical disability and cognition were explained by disconnection within specific subnetworks. In particular, higher EDSS was associated with greater structural disconnection centered around the thalami and frontal cortices, confirming previously reported relations between physical disability and disruption of fronto-thalamic and frontal commissural pathways.^11,44,49–51^ Similarly, worse performances at the SDMT were mainly explained by structural disconnection involving fronto-thalamic and frontal commissural tracts, but also long-range occipito-frontal and temporo-frontal association tracts, mostly in the left hemisphere. The relevance of thalamo-cortical, commissural and long-range association tracts for cognitive functioning, as well as the relationship between their disruption and the periventricular distribution of lesions, has been highlighted with various approaches looking at lesion location or WM microstructural properties.^42,45,52,53^ Our results confirm that atlas-based lesion disconnectomics is also sensitive to these effects, potentially providing additional information compared to the assessment of lesions and regional tissue properties.^13,54^

Morphometric similarity networks were also sensitive to MS-related brain damage, with an anatomically distributed subnetwork of similarity disruption in pwMS at baseline. The brain’s intrinsic structural organizing principles result in remote regions sharing comparable macro-scale morphological traits, thereby establishing a network of morphological similarity that can be imaged using structural MRI.^16^ Using the MIND approach, we showed that the occurrence of a relatively disordered phenomenon like MS-related neurodegeneration can disrupt this genetically determined organization, resulting in an overall reduction of morphological similarity between brain regions. These results confirm previous evidence of a more random organization of single-subject GM networks in pwMS,^17,18^ with the advantage of a method that measures multiple morphological properties simultaneously and natively aligns with macro-scale brain parcellations, therefore being more neurobiologically grounded. Morphometric similarity disruption centered around the left perisylvian cortex explained physical disability beyond whole-brain atrophy, which might speculatively be interpreted in light of the reported complex and clinically relevant anatomofunctional alterations of attentional networks in MS.^55,56^

For longitudinal network changes, we identified a subnetwork of progressive structural disconnection mainly comprising fronto-thalamic tracts, explaining disability worsening independently from lesion accrual. These results further highlight the role of cortico-thalamic connections in the pathophysiology of MS,^50^ confirming that the assessment of structural disconnection at the network level yields clinically relevant information that extends beyond mere lesion burden. Likewise, morphometric similarity networks were sensitive to longitudinal changes in the brain’s structure. Along with edges of longitudinally decreasing morphological similarity, we found pairs of regions whose morphological traits tended to match over time, substantially aligning with previously described atrophy patterns encompassing the middle temporal gyrus and sensorimotor cortices,^57^ as well as insular and prefrontal cortices and the occipital pole.^58^ Indeed, despite being a relatively disordered phenomenon, MS-related neurodegeneration is not completely random, with different spatial patterns of atrophy that have been described in association with MS.^57,58^ This disease-related structural covariance between brain regions, which is likely constrained by network-based mechanisms and shared vulnerability,^59^ may explain the observed increase in morphometric similarity. Interestingly, the longitudinal changes in morphometric similarity paralleled disability worsening independently from whole-brain volume loss, confirming its ability to capture additional information.

We also assessed the prognostic value of the assessed networks. Previous attempts have been made to predict individual-level prognosis based on lesion location,^46^ or regional network measures.^13,19^ Using NBS-predict, we demonstrated that individual structural disconnectomes can be used to significantly predict long-term disability progression independently from global lesion burden. In particular, structural disconnections of the thalamus and the paracentral lobule were the most important predictors of disability worsening, with a slight predilection for the left hemisphere. While this confirms the major role of thalamic disconnection in sustaining MS-related physical disability,^11^ the association between long-term disability progression and structural disconnection of the paracentral lobule, including the primary sensorimotor areas of the lower limbs, may be related to the known heavy dependence of EDSS on motor function and walking ability.^60^

Finally, we investigated the relationship between the two explored network domains. As expected, patients with higher global structural disconnection were also the ones with more globally disrupted morphometric similarity. On the other hand, at more granular levels, structural disconnection was associated, on average, with higher morphometric similarity. In this regard, while the distribution of values at the edge and node levels suggested a nonlinear, multiphasic relationship between structural disconnection and morphometric similarity, the positive coupling between the two might confirm the role of disconnection in shaping the patterns of concerted neurodegeneration across GM regions, with lesion-related transneuronal degeneration likely inducing similar atrophic changes at both ends of the disrupted WM tract.^59^ Our study has some limitations. First, while the assessment of structural disconnection using atlas-based approaches has been previously validated and has the advantage of greater accessibility, diffusion MRI-based tractography remains the gold standard in this regard and would have given further strength to our results. Also, the MIND approach in its standard form fails to consider subcortical GM, which is known to be highly relevant in MS, prompting the development of methodological extensions to incorporate deep GM structures in the analysis. Assessing structural disconnection and morphometric similarity changes in relation to finer clinical outcomes would help identify more focused and domain-specific network alterations, potentially informing treatment targeting. Finally, additional research relying on advanced statistical methods and prospective designs will be necessary to establish any causal relationships between the reported changes and to model their potential patterns of progression throughout the disease course.

In conclusion, our results show that networks of structural disconnection and morphometric similarity obtained from conventional MRI are sensitive to MS-related brain damage and its progression over time, potentially providing complementary information to other established MRI-derived biomarkers of disease severity and progression. Extracting network measures from conventional MRI scans holds the potential for bridging the gap between connectomics and clinical practice, driving advanced network analyses toward real-world applicability.

## Data availability

Derived data that support the findings of this study are available from the corresponding author upon reasonable request. Raw data are not available due to reasons of sensitivity.

## Acknowledgements

We thank the study participants.

