## Supplementary Figures for "Mapping Structural Disconnection and Morphometric Similarity Alterations in Multiple Sclerosis"

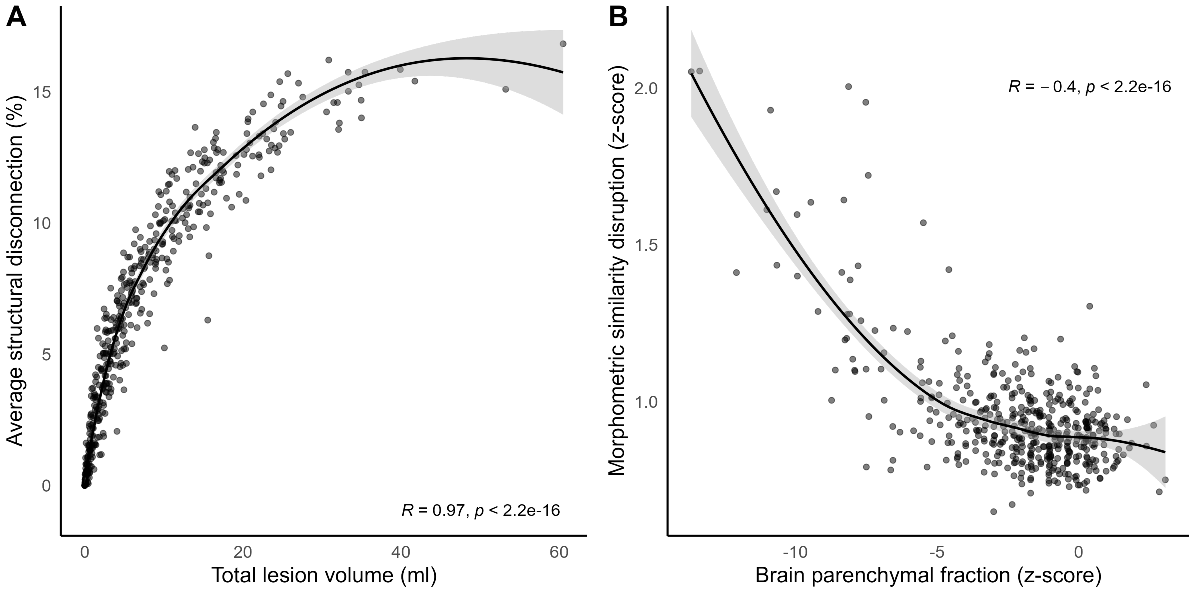


**Supplementary Figure 1.** Scatterplots showing, in pwMS, the relationship between (A) total lesion volume and average structural disconnection, and (B) brain parenchymal fraction and global morphometric similarity disruption. Local regression fit lines are shown as solid lines (with corresponding 95% confidence intervals in grey).

*Abbreviations: pwMS=patients with multiple sclerosis.*

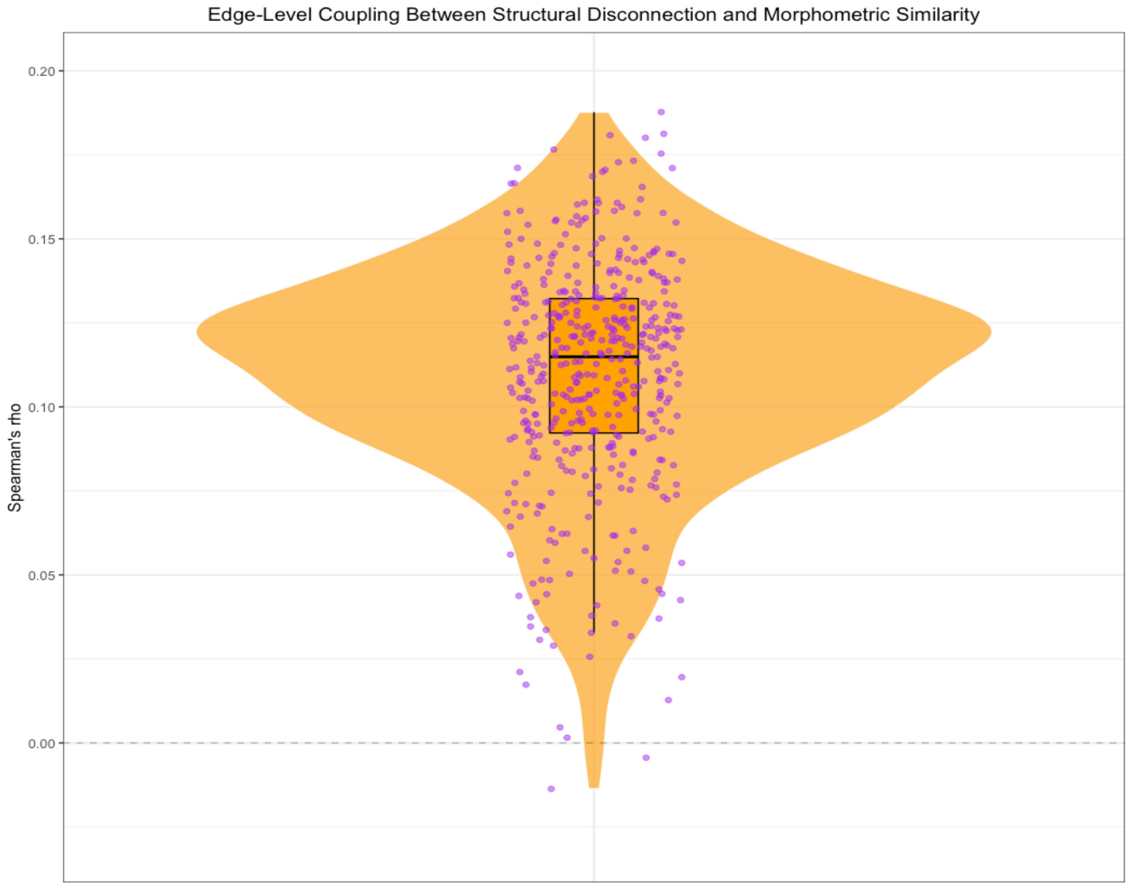


**Supplementary Figure 2.** Distribution of edge-level correlations between structural disconnection and morphometric similarity across subjects.
